# SARS-CoV-2 RNA levels in Scotland’s wastewater

**DOI:** 10.1101/2022.06.08.22276093

**Authors:** Livia C. T. Scorza, Graeme J. Cameron, Roisin Murray-Williams, David Findlay, Julie Bolland, Brindusa Cerghizan, Kirsty Campbell, David Thomson, Alexander Corbishley, David Gally, Stephen Fitzgerald, Alison Low, Sean McAteer, Adrian M. I. Roberts, Zhou Fang, Claus-Dieter Mayer, Anastasia Frantsuzova, Sumy V. Baby, Tomasz Zieliński, Andrew J. Millar

**Author notes:** corresponding author: Andrew J. Millar. these authors contributed equally to this manuscript (Livia C. T. Scorza, Graeme J. Cameron).

## Abstract

Nationwide, wastewater-based monitoring was newly established in Scotland to track the levels of SARS-CoV-2 viral RNA shed into the sewage network, during the COVID-19 pandemic. We present a curated, reference data set produced by this national programme, from May 2020 to February 2022. Viral levels were analysed by RT-qPCR assays of the N1 gene, on RNA extracted from wastewater sampled at 122 locations. Locations were sampled up to four times per week, typically once or twice per week, and in response to local needs. We report sampling site locations with geographical coordinates, the total population in the catchment for each site, and the information necessary for data normalisation, such as the incoming wastewater flow values and ammonia concentration, when these were available. The methodology for viral quantification and data analysis is briefly described, with links to detailed protocols online. These wastewater data are contributing to estimates of disease prevalence and the viral reproduction number (R) in Scotland and in the UK.

## Background & Summary

The novel coronavirus disease (COVID-19) pandemic has greatly impacted public health worldwide and has forced governments to respond quickly and implement novel strategies to surveil COVID-19 cases and closely monitor the spread of the etiological agent, the SARS-CoV-2 virus^1^.

The use of diagnostic tests is essential for disease monitoring and for the control of COVID-19 outbreaks, as it allows public health authorities to manage infected individuals using test-trace approaches, for example^2^. Several countries have adopted strategies to monitor COVID-19 cases that included reverse-transcriptase polymerase chain reaction (RT-PCR) testing together with wide availability of rapid lateral flow antigen self-tests. However, some setbacks and biases are expected in community testing. For example, a large proportion of RT-PCR tests, which are used to monitor case data, are available only for symptomatic people. Additionally, antigen tests can display a large proportion of false negatives, especially in asymptomatic individuals, so they have mostly served as an adjunct to RT-PCR tests^3^. Community testing relies on the population to volunteer and depends on the ready availability of tests. Even in countries that have resources for mass testing, the population in more remote areas might remain untested.

A complementary way of monitoring viral diseases is to use a wastewater-based epidemiology (WBE) approach. In this approach, certain viruses or drugs released by the population into the sewage network can be quantified and monitored through time ^4,5^. It has been shown that individuals infected with SARS-CoV-2 will excrete a significant number of viral particles in stools^6–8^. It was later shown that SARS-CoV-2 can be detected and quantified in wastewater samples using RT-PCR techniques or next generation sequencing and that the amount of virus in sewage correlated well with the COVID-19 case data obtained from community testing ^9–12^. This is especially useful because sewage surveillance can monitor the extent of the virus spread in the community, independent of voluntary testing, and it has the potential to detect disease outbreaks early ^9,13,14^. Several countries have used a WBE approach to monitor SARS-CoV-2, including more than 2500 sampling locations in total^15^.

The dataset presented here includes the quantification of the SARS-CoV-2 nucleocapsid gene N1 in wastewater in Scotland, UK, together with data that are complementary to the analysis of the viral concentration in each area. The SARS-CoV-2 monitoring programme in Scotland has been conducted by Scottish Water and the Scottish Environmental Protection Agency (SEPA) since May 2020. The data are released in near-real time on the SEPA public dashboard ^16^. Further statistical analysis and aggregation is performed by Biomathematics and Statistics Scotland (BioSS) and presented in weekly reports from Scottish Government ^17^. However, access to data presented in this way is at risk of deteriorating over time. It is then imperative to guarantee long-term preservation of such data and metadata whilst ensuring they adhere to FAIR principles (Findable, Accessible, Interoperable and Reusable)^18^.

The dataset and related methodologies presented here are a result of a collective effort in curating and securing future access to the outputs of the SARS-CoV-2 monitoring programme in Scotland, while ensuring they are ready to be re-used.

In this dataset, additional to the quantification of the N1 gene, we included site locations with geographical coordinates, the total population for each site, and the information necessary for data normalisation, such as the incoming wastewater flow values and ammonia concentration, when these were available. The methodology for viral quantification and data analysis is briefly described here, with links to detailed protocols on the online platform protocols.io.

## Methods

The Figure 1 summarizes the overall workflow of the SARS-CoV-2 quantification in wastewater methodology, from sample collection to data analysis and data sharing.

**Figure 1.**
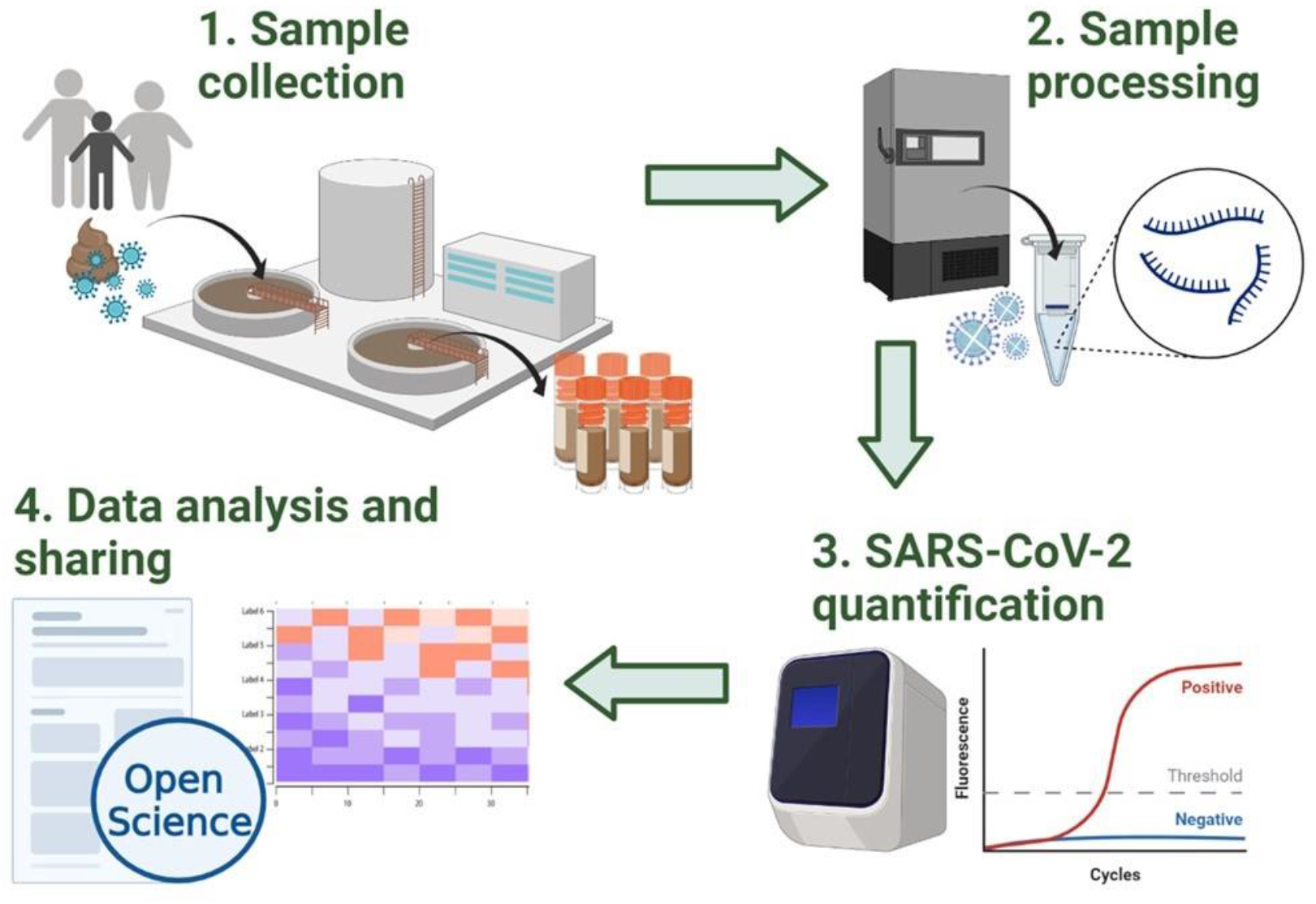
Overview of the methodology workflow of the Scottish SARS-CoV-2 monitoring in wastewater programme. Incoming wastewater samples are collected by Scottish Water (1) and transferred to SEPA’s laboratories where samples are stored and processed (2). SARS-CoV-2 viral levels are then quantified using RT-qPCR (3). Finally, the data obtained is shared on SEPA’s public dashboard and with local authorities such as the Scottish Government. Additionally, the detailed methodology and datasets are shared in open online platforms and repositories such as protocols.io, Zenodo and GitHub ^19–21,26,27^. This figure was created with BioRender.com.

The detailed methodology, from wastewater viral RNA isolation to SARS-CoV2 detection using RT-qPCR and data analysis, is described in the protocols published on the online protocol sharing platform protocols.io ^19–21^.

A brief description of the methodology for sample collection and quantification of SARS-CoV-2 in wastewater is presented here as follows.

### Sample collection

The wastewater samples were collected by Scottish Water (SW) and its operators in a total of 122 sites including all 14 NHS Scotland Health Board areas (Figure 2).

**Figure 2.**
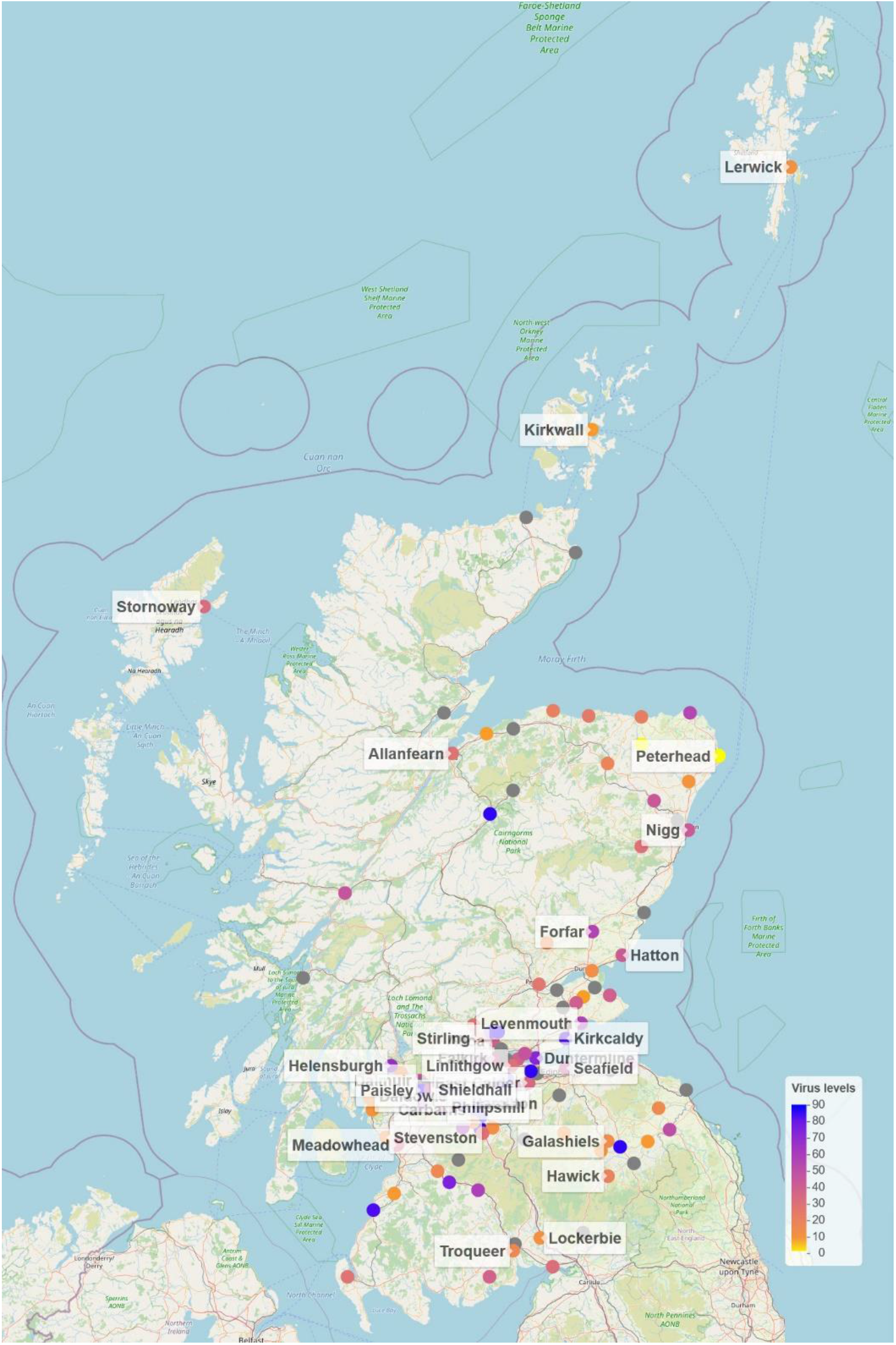
Map of Scotland showing sites from which wastewater samples were collected for SARS-CoV-2 analysis. The coloured circles show normalised SARS-CoV-2 virus levels on the last week of July 2021 in Million gene copies per person per day ([Mgc/pD]; truncated to 90). The grey circles represent sites without measurement on that week. See Methods – *Data visualization* for further details.

Samples were collected from sewage influent using either autosamplers over a period of 24 hours or, in some cases, by means of a grab sample via manholes in samples collected in other locations of the sewage network.

For samples taken at the sewage works influent, an empty bottle was put into the autosampler, and a composite sample was built up over a period of 24 hours where many small portions of the influent were taken (Day 0). Once this time period was up the sample was collected (Day 1) and transported to the SW facility. For network samples collected from a manhole as a grab sample there was no “Day 0” as the sample was directly transferred to the SW facility.

The collected samples were then split in two parts; one part was analysed for its ammonia content while the other part was transferred to SEPA’s laboratories in the next working day for SARS-CoV-2 quantification analysis (Day 2). When the number of samples was too high to be analysed at once, the samples were stored and analysed at a later date. The collection and analysis dates were recorded in the dataset (see Data Records section).

The frequency of sample collection was variable, but typically samples were collected on a weekly basis, once or twice a week. During outbreaks, for example, sample collection was more frequent in some of the health board areas.

Generally, the samples were analysed for SARS-CoV-2 RNA concentration on the day samples were received at SEPA, and the results were reported on SEPA’s public dashboard ^16^ on that same evening or the next morning.

### Sample analysis: viral RNA extraction and RT-qPCR

The majority of the samples were processed and analysed at SEPA’s laboratories whereas a few samples in January 2021 were processed and analysed by the Roslin Institute, University of Edinburgh.

Prior to processing, the samples were spiked with a known quantity of Porcine Reproductive and Respiratory Syndrome (PRRS) virus as a sample process quality control (see more details on this in the Technical Validation section). Samples were clarified to remove particles and filter-concentrated to obtain sufficient viral RNA. RNA was then extracted, and SARS-CoV-2 gene copies (as well as the PRRS control) were quantified using one-step RT-qPCR. More specifically, the amplification of the SARS-CoV-2 nucleocapsid gene N1 was monitored. After obtaining the threshold cycle (Ct) values, the gene copies per litre (gc/L) values could be calculated for each sample. Three or two technical replicates were included for each sample and the mean values were obtained for each sample. In general, the final reported value represented the mean of all replicates, but in some cases a specific replicate was deemed an outlier and excluded from the average.

### Data analysis and description

Depending on the amount of gene copies per litre obtained, samples were categorized as “Negative”, “Weak Positive”, “Positive Detected, Not Quantifiable” (“Positive DNQ”) or “Positive”. The cut-off values used to apply this description were determined by a standard dilution series analysis, which allowed the identification of the limit of detection (LoD) and the limit of quantification (LoQ) values. The LoD is the value at which the test has been determined to detect the virus material with certainty, and in this case it is currently set to be 1,316 gc/L. The LoQ is the value above which the test has been deemed to measure the virus material with a high degree of accuracy, and in this case it is currently set at 11,368 gc/L. The category thresholds were slightly altered from one point within the reported interval to reflect a change in lab practice. These instances can be identified by the category “SEPA-Low Volume” in the “Analysis_Lab” column of the dataset. In those particular instances, the analysis volume was reduced, which reflected in changes in the LoD and LoQ values. When this change happened, the LoD increased to 6,640 gc/L and the LoQ reduced to 9,427 gc/L.

If two or more replicates produced no Ct, the result was reported as “Negative” and the N1 reported value was set to zero. If two out of three replicates or one out of two replicates returned a positive signal, but that signal was calculated as lower than the LoD, the sample was reported as “Weak Positive. If the average value of the three or two replicates was between the LoD and the LoQ (i.e. between 1,316 and 11,386 gc/L), the sample was reported as “Positive (DNQ)” as it could not be quantified in a statistically significant way. Finally, a “Positive” description was given where the average of all replicates was above the LoQ and therefore showed a strong, quantifiable result.

In some rare instances where the analysis could not be done due to technical issues, the sample was described as “Analysis failed” and the cells which should contain the N1 reported values are blank.

### Normalisation

Although the gene copies per litre values described above are useful indicators of the presence of the virus in each specific site, they do not consider the dilution factor caused by the incoming flow in wastewater or the size of the population of each site. For example, in periods of heavy rainfall, the viral particles will be more diluted in the wastewater compared with periods with less rain. Additionally, in some areas the industrial discharges in wastewater will affect the incoming flow and therefore the viral concentration coming from household waste. The size of the population can also produce biases. A certain number of positive individuals in a small catchment area can produce higher values of viral gene copies compared with the same number of positive individuals in a larger catchment area.

Therefore, a better representation of the spread of the virus within the population can be derived from normalised values that use the site population and the incoming wastewater flow to determine the million gene copies per person per day [Mgc/pD] ^22,23^. The detailed normalisation protocol is provided on protocols.io ^21^. In short, to produce a daily value of million gene copies per person, the raw gene copies per litre value is multiplied by the daily flow total and divided by the population served at each site. The flow for a specific site (waterworks location) is either measured directly or estimated using an estimation method that will vary according to the availability of the supporting data. For example, if ammonia data are available, this can be used to derive the flow values. Flow values can also be derived from an average of the historical flow data for the site, when these are available.

### Data visualization

The figure with geographical locations of the sites (Figure 2) was created with R using the *leaflet* package ^24^. The circle markers representing each site location were colorized according to the averaged normalised SARS-CoV-2 virus levels on the last week of July 2021. The colour scale was limited to 90 [Mgc/pD] to highlight differences between virus levels, and the sites without measurements on that week were marked in black. For clarity, only a few selected sites for each health board were labelled with their names.

The heatmap (Figure 3) was created with R using the *ComplexHeatmap* package ^25^. To obtain a heatmap showing weekly values for SARS-CoV-2 gene copies per person per day, the normalised gene copies values falling into the same week for each site were averaged.

**Figure 3.**
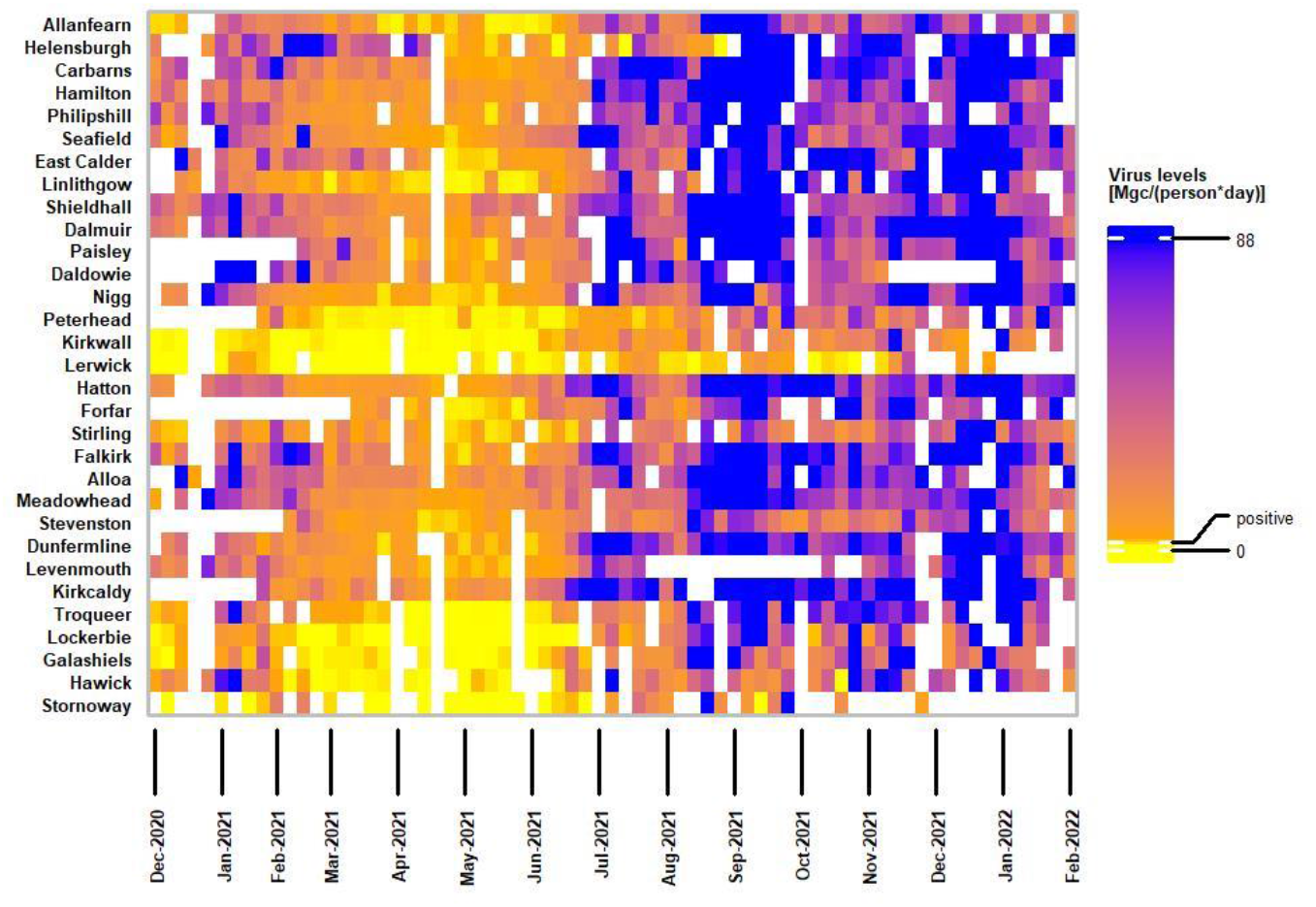
Heatmap showing SARS-CoV-2 RNA levels over time at selected sampling sites, illustrating key features of the data across the Alpha, Delta and Omicron waves. The sites represent all 14 Scotland NHS health boards. To obtain the weekly normalized values, the gene copies per person per day falling into the same week for each site were averaged. To build the heatmap, two break points were used in the visualisation colour scale: 2.9 (labelled positive) and 88, which correspond to 3rd quartiles of normalized data labelled as non-positive and positives, respectively. See Methods – *Data visualization* for further details.

To build the heatmap, three break points were used in the visualisation colour scale: 0, 2.9 (labelled positive), and 88. To get to these values, the normalised virus levels were filtered according to the criteria that characterised them as positives and non-positive (i.e. negative, weak positive and positive DNQ), 2.9 is 3^rd^ quartile of all non-positive values, 88 is 3^rd^ quartile of the positive values range. The quartiles were used rather than min/max as the values are long tailed and the normalization introduces additional outliers, for example positive values span to over 23000 millions copies per person per day.

The detailed R code for figures preparation can be found on our GitHub page ^26^.

### Data Records

The data presented here is available on a GitHub repository ^26^ and on Zenodo ^27^. The datasets are provided in csv format.

The main dataset represents data obtained from the quantification of the N1 gene of SARS-Cov-2 in wastewater in all 14 NHS Scotland Health Board areas, comprising a total of 122 sampling sites. A description of each column within the file named “SARS-Cov2_RNA_monitoring_ww_scotland.csv” is provided below:

- **Health_Board**: name of the NHS Scotland health board for that particular site and sample.
- **Site**: name of the site where the sample was collected, which corresponds to the name of the wastewater treatment centre. In some cases, the site name is followed by a dash and a specific sewage location within the main treatment centre network. Ex. “Seafield -Western General”.
- **Date_collected**: date at which the wastewater sample was collected.
- **Date_analysed**: date at which the sample was analysed. In some instances, cells in this column contain “(empty)” because the exact date of the analysis was not available.
- **SW_sample_number**: Scottish Water (SW) sample number. The majority of the samples collected by SW have a sample number assigned. The numbers were assigned in parallel with shipment to the SEPA lab, hence in rare instances the sample arrived but its number did not.
- **N1_description**: category assigned to the result from analysis of the sample by RT-qPCR. This value can be “Negative”, “Weak Positive”, “Positive (DNQ)”, “Positive” or “Analysis Failed”. See *data analysis and description* on Methods for more details.
- **N1_Reported_value-gc_per_L**: the gc/L value reported on SEPA’s public dashboard. These reported values are normally the calculated mean of all replicates. In some cases, the reported value excluded an outlier replicate. The value is set to zero when the replicates did not produce a Ct value or the mean was below the LoD. See *data analysis and description* on Methods for more details.
- **N1_Repl_1-gc_per_L**: the gc/L of replicate 1. Blank fields (null values) mean that a technical issue occurred, and a result was not produced for that sample. The value was set to zero when the replicate did not produce a Ct value.
- **N1_Repl_2-gc_per_L:** the gc/L of replicate 2. Blank fields (null values) mean that a technical issue occurred, and a result was not produced for that sample. The value was set to zero when the replicate did not produce a Ct value.
- **N1_Repl_3-gc_per_L:** the gc/L of replicate 3. Blank fields (null values) mean that a technical issue occurred, and a result was not produced for that sample. Blank fields (null values) in this case might also mean that a third replicate was not included for that sample. The value was set to zero when the replicate did not produce a Ct value.
- **Calculated_mean**: the simple mean of the values obtained for all replicates.
- **Standard_Deviation**: the standard deviation of the values obtained for all replicates.
- **Flow-L_per_day**: the measured flow in litres per day for that particular site and sample collection date. As described in the methodology, not all samples have a flow measurement associated.
- **Ammonia-mg_per_L**: the measured ammonia content in milligrams per litre for that particular site and sample collection date. As described in the methodology, not all samples have an ammonia measurement associated.
- **pH_value**: the pH of the sample, when available.
- **Modelled_flow-L_per_day:** the modelled flow in litre per day produced according to the methodology described in the normalisation process. This was particularly used when flow measurements were not available for a specific site and date.
- **Million_gene_copies_per_person_per_day**: to produce these values, the raw gc/L measurements were multiplied by the daily flow total (or modelled flow), and divided by the population served at each site, to produce a daily value of RNA copies per person. The detailed methodology is described in the methods section and on the detailed protocol on protocols.io ^21^.
- **Analysis_lab**: the laboratory in which the samples were analysed. In most cases the samples were analysed by SEPA, but in some instances, samples were analysed by the The Roslin Institute, University of Edinburgh. The “SEPA-Low Volume” is an identification for a change in laboratory process that occurred in a period of time. This description means that in that particular analysis the volume was reduced, which reflected in changes in the LoD and LoQ values. See *data analysis and description* on Methods for more details.

To facilitate re-use, we prepared transformed/secondary data based on the main data file. The file “SARS-Cov2_RNA_monitoring_ww_scotland_full.csv” contains the de-normalise data with the additional information about the collection sites inserted, like their geographic locations and population in the capture area. The file contains the same columns as SARS-Cov2_RNA_monitoring_ww_scotland.csv plus:

- Latitude_dd – geographic position of the sample collection sites (north/south of the equator) in decimal degrees.
- Longitude_dd – geographic position of the sample collection site (east/west of the meridian) in decimal degrees.
- Population – number of people in the sewage catchment upstream of the sampling site.
- Population_band – size of the population in one of the following bands: 0-2K, 2K-4K, 4K-10K, 10K-100K, 100K+.

The file “prevalence_timeseries.csv” contains data in the traditional timeseries format, i.e., each row corresponds to one sample and collection site, the first few columns provide information about the site, and the subsequent columns store SARS-CoV-2 virus levels measurements for each particular sampling date. For example, the column “2020-05-28” contains gene copies per litre from sample collection on 2020-05-28. NA means that a sample was not collected for that site at that date or that the analysis failed.

The file “norm_prevalence_timeseries.csv” is equivalent to the “prevalence_timeseries.csv” but the “date” columns contain the normalised virus levels in million gene copiers per person per day.

The file “weekly_prevalence_timeseries.csv” – contains weekly averaged data of virus levels in the timeseries format, i.e., each row corresponds to one sample and collection site, the first few columns provide information about the site, and the subsequent columns store averaged SARS-CoV-2 virus levels recorded for a week. For example, 2022-6 contains averaged gene copies per litre from samples collected on the week starting at 2022-02-7 (sixth week of the year, 2^nd^ week of February). NA means that no samples were collected in that week for a site or that all the analysis of that week failed.

Finally, the file “weekly_norm_prevalence_timeseries.csv” – is equivalent to the “weekly_ prevalence_timeseries.csv” but the “week” columns contain the averaged normalized virus levels in million gene copies per person per day for that week.

As described in the methodology, it took a maximum of two working days from sample collection to reporting the SARS-CoV-2 quantification results on SEPA’s public dashboard ^16^. During the total period of monitoring described in this dataset (May 2020 to February 2022), 9.3% of the analysed samples were “Negative”, 7.3% were “Weak Positives”, 14% were “Positive DNQs”, and 63.4% were “Positive”. The analysis failed in 6% of the samples. Figure 3 shows normalised data (million gene copies per person per day) from December 2020 to February 2022 obtained from 31 selected sites representing all 14 health boards. This includes the sites with highest population such as Seafield (Edinburgh), Shieldhall (Glasgow) and Nigg (Aberdeen) as well as remote sites such as Kirkwall and Lerwick (Orkney and Shetland; Northern Isles) and Stornoway (Western Isles). The normalised data, which describes the number of million gene copies per person per day shows that the wastewater data correlates well with the COVID-19 disease waves reported by local authorities ^28^ (Fig. 3). Nationally, an increase in cases was observed during the period of December 2020 to February 2021, followed by a period with fewer cases from February 2021 until mid-June 2021, when cases started to rise again. This is observable also in wastewater data, where we can visualize the same pattern in the levels of SARS-CoV-2 viral material (Fig. 3). Moreover, in September 2021 a higher peak in case data correlated with higher number of viral copies in wastewater data, followed by a slight decrease in cases before another peak was observed from end of December 2021 until February 2022.

The pH was measured only in a small proportion of the samples (approximately 9%) in 57 of the total number of sites. The pH values were not recorded from April 2021, as these values did not impact the analysis.

Additional to the data related to the SARS-CoV-2 N1 gene quantification, data related to the population in the sewage catchment upstream of each sampling site and the site’s geographical coordinates are also provided.

### Technical Validation

A quality control was included in each measurement to detect any technical issues with viral RNA extraction, cDNA synthesis and PCR amplification. This procedure involved adding a known quantity of a non-target RNA to each wastewater sample prior to sample processing. The Porcine Reproductive and Respiratory Syndrome (PRRS) virus was used as the “spike-in” control and specific primers for this virus were used during the RT-qPCR analysis. The successful detection of material from the PRRS virus demonstrated that the methodology to concentrate, isolate and recover SARS-CoV-2 RNA was successful and that any negative results were due to the absence of the gene of interest (N1) in the sample and not because of methodological failures. Similarly, an extraction blank was included to help with the detection of any contamination. For RT-qPCR experiments three or two technical replicates were included for each sample, as described above.

Additionally, serial dilutions of template RNA were used to produce standard curves and therefore establish the experimental efficiency and reliability and allow for the absolute quantification of the target gene.

### Usage Notes

Mathematical models using wastewater data can be used as a guide to estimate daily cases of COVID-19. For example, modeling based on wastewater monitoring data has been used in Scottish Government to estimate the prevalence of infection, and the value of R^17^.

Wastewater data therefore provides estimates of these values that are independent of other data types. These estimates are combined with others to provide the best overall estimate of prevalence and R values, at the Scottish and UK levels.

The detailed methodology accompanying this dataset, including normalisation methods, can assist with modelling and predicting surges in SARS-CoV-2 or in similar viral diseases in the future. The methodology for SARS-CoV-2 detection presented here can also be adopted by other institutions interested in WWE monitoring or modified to monitor different viruses. Moreover, longitudinal, geospatial data are costly to obtain – especially taking into consideration the logistics of collecting physical samples – and thus, warrant appropriate preservation strategies.

Transparency of data regarding the COVID-19 epidemic is crucial when it comes to government decision making and accountability. For example, availability of such data can potentially help researchers trace back the viral levels in the community and relate them to government actions and the following chain of events that took place. Additionally, knowing that COVID-19 is a new disease but with already more than fifty long-term effects ^29^ and potentially many more, this data has the potential to help researchers and medical professionals to correlate and understand surges in certain medical conditions in areas that were particularly affected by the virus.

### Code Availability

All codes produced by this work for data analysis are available in the GitHub repository ^26^with rights to access under the Creative Commons (CC) BY License.

The software environment used for data analysis including data normalisation and the heatmap and map figures (Figs. 2-3) was R.

## Data Availability

All data produced are available online in the data repositories cited in the manuscript

https://doi.org/10.5281/zenodo.6339631

https://github.com/BioRDM/COVID-Wastewater-Scotland

## Acknowledgements

We thank Scottish Water for providing all samples used in this project. This work was funded by Scotland’s Centre of Expertise for Waters (CREW; Grant CD2019_06 Tracking SARS-CoV-2 via municipal wastewater) and by Scottish Environment Protection Agency (SEPA).

## Author contributions

LCTS took the lead in writing the manuscript with input from all authors. GC and RMW analysed, curated and distributed the data. DF, JB, BC, KC, DT developed the RNA extraction and RT-qPCR protocols presented here and produced the data related to quantification of the SARS-CoV-2 N1 gene in wastewater. DF supervised the implementation of the aforementioned methods. AMIR, ZF, CDM, AF developed the normalisation methodology. AC, DG, SF, AL, SM developed the first methodologies of the Covid-19 WBE Scotland programme, which substantiated this work. LCTS, TZ, SB curated the dataset for this paper and produced the figures of this manuscript. TZ and AJM supervised the data curation process. AJM contributed to the management of the COVID-19 WBE programme for Scottish Government.

